# Discordant humoral and T cell immune responses to SARS-CoV-2 vaccination in people with multiple sclerosis on anti-CD20 therapy

**DOI:** 10.1101/2021.08.23.21262472

**Authors:** Sachin P. Gadani, Maria Reyes-Mantilla, Larissa Jank, Samantha Harris, Morgan Douglas, Matthew D. Smith, Peter A. Calabresi, Ellen M. Mowry, Kathryn C. Fitzgerald, Pavan Bhargava

**Affiliations:** Division of Neuroimmunology, Department of Neurology, Johns Hopkins University School of Medicine, Baltimore, MD, USA

**Author notes:** Corresponding Authors: Pavan Bhargava, MD, Department of Neurology, 600 N Wolfe St, Pathology 627, Baltimore, MD 21287., Kathryn Fitzgerald, ScD, Department of Neurology, 600 N Wolfe St, Pathology 627, Baltimore, MD 21287. These authors contributed equally (Sachin P. Gadani, Maria Reyes-Mantilla). Senior Authors (Kathryn C. Fitzgerald, Pavan Bhargava).

## Abstract

**Background:** Sphingosine-1-phosphate receptor (S1P) modulators and antiCD20 therapies impair humoral responses to SARS-CoV-2 mRNA vaccines. Whether disease modifying therapies (DMTs) for multiple sclerosis (MS) also impact T cell immune response to vaccination is unknown.

**Methods:** In 101 people with MS, we measured humoral responses via an immunoassay to measure IgG against the COVID-19 spike S1 glycoprotein in serum. We also measured T cell responses using FluoroSpot assay for interferon gamma (IFN-γ) (Mabtech,Sweden) using cryopreserved rested PBMCs and then incubated in cRPMI with 1µg/ml of pooled peptides spanning the entire spike glycoprotein (Genscript, 2 pools; 158 peptides each). Plates were read on an AID iSpot Spectrum to determine number of spot forming cells (SFC)/10^6^ PBMCs. We tested for differences in immune responses across DMTs using linear models.

**Findings:** Humoral responses were detected in 22/39 (56.4%) participants on anti-CD20 and in 59/63 (93.6%) participants on no or other DMTs. In a subset with immune cell phenotyping (n=88; 87%), T cell responses were detected in 76/88 (86%), including 32/33 (96.9%) participants on anti-CD20 therapies. AntiCD20 therapies were associated with an increase in IFN-γ SFC counts relative to those on no DMT or other DMTs (for antiCD20 vs. no DMT: 425.9% higher [95%CI: 109.6%, 1206.6%] higher; p<0.001; for antiCD20 vs. other DMTs: 289.6% [95%CI: 85.9%, 716.6%] higher; p<0.001).

**Interpretation:** We identified a robust T cell response in individuals on anti-CD20 therapies despite a reduced humoral response to SARS-CoV-2 vaccination. Follow up studies are needed to determine if this translates to protection against COVID-19 infection.

## Introduction

The COVID-19 pandemic has impacted people with multiple sclerosis (MS), both directly, as a result of morbidity and mortality from COVID-19 as well as indirectly through uncertainty in how best to optimize MS care during this time^1^. For example, certain disease modifying therapies (DMTs) may impact the risk of contracting COVID-19 or developing severe COVID-19 infection,^2,3^ and, it is unclear whether certain DMTs should be held or modified in how they are used^3^.

The introduction of highly effective vaccines, such as the SARS-CoV-2 mRNA vaccines produced by Pfizer and Moderna, provides an effective intervention to reduce the risk and severity of COVID-19 infection^4,5^. Multiple studies indicate that COVID-19 vaccination results in both a humoral and cell-mediated immune response that is likely to play a role in their protective effects^6^.

Certain MS DMTs such as anti-CD20 therapies or sphingosine-1-phosphate (S1P)-receptor modulators can impact responses to a variety of existing vaccines,^7,8^ and emerging studies suggest these therapies may also impair humoral response to SARS CoV-2 vaccines^9,10^ Prior studies also suggest the T cell immune response may be maintained following administration of other common vaccines in patients treated with anti-CD20 therapies; however, to our knowledge, studies have not yet assessed cell-mediated immune responses to SARS-CoV-2 vaccination.^11^ Herein, we assessed both humoral and T cell responses to vaccination in people with MS on a range of DMTs.

## Methods

### Recruitment and Sample Collection

Blood was collected from donors following full informed consent under a protocol approved by the Johns Hopkins Medicine Institutional Review Board. We recruited patients with Multiple Sclerosis on a volunteer basis from the Johns Hopkins MS center who had recently received the COVID19 vaccine and were part of the COVID-RIMS study^3^. No patients were excluded from the study based on type of disease modifying therapy, COVID19 vaccine (all patients received either Pfizer, Moderna, or Johnson & Johnson vaccines), or any other demographic or disease characteristic. Recruited patients underwent phlebotomy either 4 or 8 weeks after the terminal COVID19 vaccination dose.

### Humoral response assay

Serum was isolated by centrifuging coagulated blood using a standardized protocol. We measured serum humoral responses using an ELISA quantifying IgG specific to the COVID-19 spike S1 glycoprotein (EUROIMMUN, Germany), which was given emergency use authorization by the Food and Drug Administration^12^. This ELISA was performed in a Clinical laboratory improvement amendments (CLIA) certified laboratory at the Johns Hopkins Department of Pathology^12^. This assay has high sensitivity and specificity and correlates with presence of neutralizing antibodies. The cut-off value for the presence of a humoral response on this assay is 1.24 and details on performance of this assay and determination of this cut-off have been reported previously^12^.

### T cell response assay

Peripheral blood mononuclear cells (PBMCs) were isolated via centrifugation in a Ficoll gradient (using SepMate PBMC isolation tubes, STEMCELL technologies) and cryopreserved in media containing 10% DMSO. PBMCs were thawed and rested for 12 hours in complete culture media (RPMI + 10% Fetal Bovine Serum). PBMCs were plated into a 96-well FluoroSpot assay plate for interferon gamma (IFN-γ) (Mabtech, Sweden, FSP-0102-10) at 2.5 × 10^5^ cells per well for stimulation. Pooled peptides spanning the length of the entire spike glycoprotein (2 pools of 158 peptides each; Genscript, RP30020) were used for stimulation at a concentration of 1ug/mL per peptide. Positive controls were stimulated with anti-CD3/anti-CD28 and negative controls received no stimulation. Three technical replicates were completed for each condition. After 22 hours of stimulation, cells were discarded and FluoroSpot plates were prepared per manufacturer’s instructions. Plates were read on an AID iSpot Spectrum in the Johns Hopkins Immune Monitoring Core lab. Results were expressed as spot forming cells (SFC)/10^6^ PBMCs and were obtained by subtracting the average counts for the negative control from the average counts for each peptide pool and then summing the counts for the two peptide pools. A negative T cell response was defined as the lack of response to both peptide pools – stimulation index (counts for the peptide pool divided by count in the negative control) of less than 3 or count of <20 SFC/10^6^ PBMCs for each peptide pool^13^.

### Statistical methods

Initial descriptive statistics assessed differences in demographic or MS characteristics across therapy groups. We categorized patients on considered those on glatiramer acetate and interferon-beta into an any injectable therapy category. We also collapsed dimethyl fumarate and teriflunomide into an oral therapies group; we did not include fingolimod in the oral therapies group because of initial findings of other groups suggesting a lack of humoral vaccine response for individuals on this therapy specifically. We assessed the association between therapy class and odds of humoral vaccine response using a logistic regression model adjusted for age, sex, and time from first dose of the vaccine to blood collection (as samples were collected variably - 4 or 8 weeks following terminal vaccine dose). A similar model assessed whether time from last infusion were associated with odds of a humoral response in individuals treated with antiCD20 therapies. We next assessed the association between therapy classes and log-transformed IFN-γ SFC adjusted for age, sex and time from first dose of the vaccine. As only 3 individuals were treated with fingolimod, we did perform formal analyses assessing differences in IFN-γ SFC counts for this therapy.

## Results

We enrolled 101 participants (82% female), 94% of whom received an mRNA vaccine (94%) with blood collection an average 6.8 weeks after terminal vaccine dose (**Table 1**).

**Table 1.** Demographic characteristics of study cohort.

|  | Disease Modifying Therapy Category |  |  |  |  |  |
| --- | --- | --- | --- | --- | --- | --- |
|  | None | Injectable | Natalizumab | Other oral | AntiCD20 | Fingolimod |
| **N** | 14 | 16 | 16 | 14 | 38 | 3 |
| **Age, years, mean (SD)** | 57.42 (12.84) | 50.17 (8.52) | 47.63 (9.01) | 49.12 (11.27) | 47.78 (9.64) | 47.93 (9.36) |
| **Male sex, n (%)** | 3 (21.4) | 3 (18.8) | 1 (6.2) | 2 (14.3) | 9 (23.7) | 0 (0.0) |
| **Race, n (%)** |  |  |  |  |  |  |
| White | 12 (85.7) | 16 (100.0) | 15 (93.8) | 14 (100.0) | 34 (89.5) | 3 (100.0) |
| Black | 0 (0.0) | 0 (0.0) | 0 (0.0) | 0 (0.0) | 3 (7.9) | 0 (0.0) |
| Other | 2 (14.3) | 0 (0.0) | 1 (6.2) | 0 (0.0) | 1 (2.6) | 0 (0.0) |
| **Hispanic or Latino ethnicity, n (%)** | 0 (0.0) | 0 (0.0) | 3 (18.8) | 0 (0.0) | 1 (2.6) | 0 (0.0) |
| **Vaccination manufacturer, n (%)** |  |  |  |  |  |  |
| J&J | 0 (0.0) | 0 (0.0) | 2 (12.5) | 2 (14.3) | 3 (7.9) | 0 (0.0) |
| Moderna | 6 (42.9) | 8 (50.0) | 5 (31.2) | 4 (28.6) | 10 (26.3) | 0 (0.0) |
| Pfizer | 8 (57.1) | 7 (43.8) | 9 (56.2) | 7 (50.0) | 23 (60.5) | 3 (100.0) |
| Unsure | 0 (0.0) | 1 (6.2) | 0 (0.0) | 1 (7.1) | 2 (5.3) | 0 (0.0) |
| **MS subtype, n (%)** |  |  |  |  |  |  |
| PPMS | 2 (14.3) | 0 (0.0) | 0 (0.0) | 0 (0.0) | 3 (7.9) | 0 (0.0) |
| RRMS | 9 (64.3) | 16 (100.0) | 14 (87.5) | 14 (100.0) | 32 (84.2) | 3 (100.0) |
| SPMS | 3 (21.4) | 0 (0.0) | 2 (12.5) | 0 (0.0) | 3 (7.9) | 0 (0.0) |
| **Individual DMT, n (%)** |  |  |  |  |  |  |
| none | 14 (100.0) | - | - | - | - | - |
| rituximab | - | - | - | - | 1 (2.6) | - |
| ocrelizumab | - | - | - | - | 37 (97.4) | - |
| dimethyl fumarate | - | - | - | 13 (92.9) | - | - |
| teriflunomide | - | - | - | 1 (7.1) | - | - |
| fingolimod | - | - | - | - | - | 3 (100.0) |
| glatiramer acetate | - | 7 (43.8) | - | - | - | - |
| interferon beta | - | 9 (56.2) | - | - | - | - |
| natalizumab | - | - | 16 (100.0) | - | - | - |
| **Disease duration, years, mean (SD)** | 9.15 (9.77) | 12.00 (6.56) | 13.07 (9.24) | 12.86 (10.75) | 9.51 (7.31) | 9.33 (7.64) |
| **DMT therapy duration, mean (SD)** | - | 10.84 (28.40) | 2.28 (2.20) | 1.43 (1.94) | 1.95 (0.88) | 3.75 (4.79) |

### Humoral response to SARS-CoV-2 vaccination

All participants on no therapy (n=14), injectables (n=16) or natalizumab (n=16) and the majority on non-S1P modulating oral therapies (12/14; 86%) demonstrated a humoral response to vaccination (Figure 1A). In contrast, only 22/39 (56%) of participants exposed to anti-CD20 therapy and 1/3 participants on S1P modulating therapy exhibited a humoral response to vaccination (Figure 1A). Among patients on anti-CD20 therapy, a 30 day increase in time from last infusion was associated with 1.45 increased odds of a positive humoral response to vaccination (**Figure 1B**; OR: 1.51; 95% CI: 1.05-2.17).

**Figure 1.**
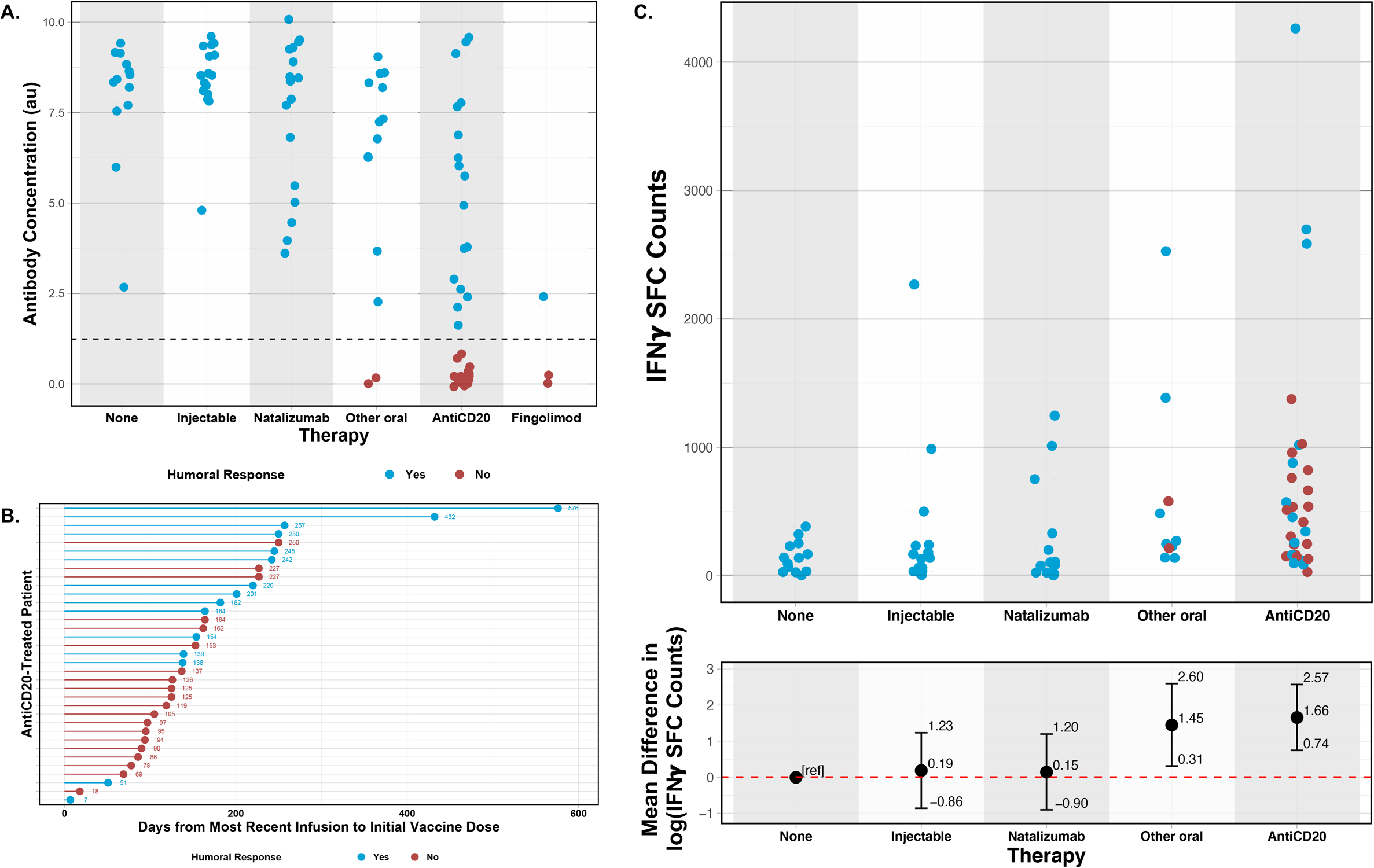
Humoral and cell mediated responses to SARS-CoV-2 vaccination in people with multiple sclerosis. **A**. Dot plot of IgG levels against S1 subunit of spike glycoprotein by disease modifying therapy (DMT) category; dotted line is cut-off for positivity of antibody response to vaccination. **B**. Time from last infusion of B-cell depleting agent in the study population; lines are color coded based on antibody response status to vaccination and displayed values note the number of days from most recent infusion to first vaccination dose; mean (SD) time from last infusion was 165 (109) days. **C**. Dot plot of T cell response to spike glycoprotein peptides (number of IFNγ producing cells/10^6^ PBMCs) by DMT category [above]. The bottom panel depicts the age and sex-adjusted mean difference in log-transformed IFNγ SFC counts relative to MS patients not on a DMT.

### T cell response to SARS-CoV-2 vaccination

Most participants (76/88, 86%) across all DMTs demonstrated a T cell immune response to SARS CoV-2 vaccination. Interestingly, participants on anti-CD20 or non-S1P modulating oral therapies had significantly higher IFNγ SFC counts compared to those on no DMT (**Figure 1C**). Participants on anti-CD20 therapy in particular had on average 1.36 higher log(IFNγ SFC counts) when compared to individuals on other DMTs (mean difference in log[IFNγ SFC counts] versus other DMTs: 1.36; 95% CI: 0.62, 2.10; p<0.001).

## Discussion

The use of immunosuppressive medications was an exclusion criteria in the phase 3 clinical trials for most SARS-CoV-2 vaccines^5^, producing a critical gap in our understanding of their safety and efficacy in patients being treated for MS and other autoimmune conditions. In this study, we found that patients treated with anti-CD20 therapy had impaired humoral response to SARS-CoV-2 vaccination, consistent with previous reports^9,10^. Interestingly, the T cell response to SARS-CoV-2 vaccination was more robust in anti-CD20 treated patients relative to patients not on a DMT or those on other DMTs, even in those anti-CD20 treated patients lacking an antibody response. We also found that most patients treated with non-anti-CD20 therapies developed a robust humoral and cellular response to SARS-CoV-2 vaccination.

B cells, in addition to producing antibodies, also present antigens and are important activators of CD4+ and CD8+ T cells^14^. Our results suggesting that B cell depletion increases T cell vaccine responses are therefore surprising and warrant further investigation. In agreement with our results a recent study showed that patients with X-linked agammaglobulinemia also mounted a stronger T cell response to SARS-CoV-2 vaccination compared to healthy controls^15^. Possible mechanisms underlying this finding include depletion of regulatory B cells, alleviating their inhibition of T cell activation^16^, decreased activation of regulatory T cells in the absence of B cells^17^, or alterations in the local inflammatory mileu at the site of vaccination.

While most patients with MS are not at significantly higher risk of morbidity and mortality from COVID-19 infection, the use of anti-CD20 therapies has been linked to greater COVID-19 severity in registry studies^2,18^. The reduction of humoral responses in these patients has raised concerns that people on anti-CD20 therapy may have a greater COVID-19 infection risk despite vaccination and has prompted discussion of booster doses, perhaps in conjunction with delaying therapy infusions, to mitigate this risk. However, since stronger SARS-CoV-2 specific T cell responses in the setting of natural infection have been linked to lower disease severity^6^, our data demonstrating robust T cell responses to SARS-CoV-2 vaccination in patients on anti-CD20 therapy suggests that vaccination likely confers some promise of protection, even in the absence of detectable humoral responses. Confirmation of this observation will require follow-up studies examining post-vaccination breakthrough infections in patients on anti-CD20 therapies.

In conclusion, this study provides novel information regarding T cell responses to SARS-CoV-2 vaccination in people with MS on a variety of DMTs, identifying robust T cell responses even in patients on anti-CD20 therapy who do not mount a humoral response to SARS-CoV-2 vaccination.

## Data Availability

De-identified data may be available upon request for investigators whose proposed use has been approved by the Institutional Review Board (IRB) of institution of the principal investigator.

## Research in context

### Evidence before this study

We searched PubMED for the term ((“COVID-19”) or (“SARS-CoV-2”) or (“coronavirus”)) AND ((“vaccination”) or (“vaccine”)) AND (“multiple sclerosis”), published between January 1, 2020 and August 20, 2021. 11 results evaluated the humoral response to SARS-CoV-2 vaccination in people with MS on a variety of disease modifying therapies and noted a reduction in humoral response to vaccination in patients on anti-CD20 and sphingosine-1-phosphate receptor modulators. However, we found no published reports describing T cell responses to SARS-CoV-2 vaccination in people with MS.

### Added value of this study

We conducted this study to address the lack of current knowledge regarding T cell responses to SARS-CoV-2 vaccination in people with MS and the effect of disease modifying therapies (DMTs) on this response. In a study involving 101 people with MS, we confirmed previous findings that the humoral response to SARS-CoV-2 vaccination was reduced in people with MS on anti-CD20 therapy. We also noted that prolonged time from last infusion of anti-CD20 therapy was associated with higher chance of a positive humoral response to SARS-CoV-2 vaccination. We found that the majority of people with MS mounted a T cell response to SARS-CoV-2 vaccination with those on anti-CD20 mounting a more robust T cell response than those not on a treatment or on other DMTs.

### Implications of all the available evidence

Use of anti-CD20 agents is associated with lower humoral response to SARS-CoV-2 vaccination in people with MS. Identification of a more robust T cell response in people with MS suggests at least partial efficacy of vaccines and some potential for protection from severe COVID-19 disease even in the absence of a humoral immune response.

## Acknowledgements

This study was funded partially by 1K01MH121582-01 from NIH/NIMH and TA-1805-31136 from the National MS Society (NMSS) to KCF and TA-1503-03465 and JF-2007-37655 from the NMSS to PB. This study was also supported through the generosity of the collective community of donors to the Johns Hopkins University School of Medicine for COVID research.

## Author Contributions

Conception and design of study – PAC, EMM, KCF and PB; Acquisition and analysis of data – SPG, MR, LJ, SH, MD, MDS, KCF and PB; Drafting a significant portion of the manuscript or figures – SPG, MR, PAC, EMM, KCF and PB.

